# Neutrophil extracellular traps (NETs) as markers of disease severity in COVID-19

**DOI:** 10.1101/2020.04.09.20059626

**Authors:** Yu Zuo, Srilakshmi Yalavarthi, Hui Shi, Kelsey Gockman, Melanie Zuo, Jacqueline A. Madison, Christopher Blair, Andrew Weber, Betsy J. Barnes, Mikala Egeblad, Robert J. Woods, Yogendra Kanthi, Jason S. Knight

## Abstract

In severe cases of coronavirus disease 2019 (**COVID-19**), viral pneumonia progresses to respiratory failure. Neutrophil extracellular traps (**NETs**) are extracellular webs of chromatin, microbicidal proteins, and oxidant enzymes that are released by neutrophils to contain infections. However, when not properly regulated, NETs have potential to propagate inflammation and microvascular thrombosis—including in the lungs of patients with acute respiratory distress syndrome. While elevated levels of blood neutrophils predict worse outcomes in COVID-19, the role of NETs has not been investigated. We now report that sera from patients with COVID-19 (n=50 patients, n=84 samples) have elevated levels of cell-free DNA, myeloperoxidase(**MPO**)-DNA, and citrullinated histone H3 (**Cit-H3**); the latter two are highly specific markers of NETs. Highlighting the potential clinical relevance of these findings, cell-free DNA strongly correlated with acute phase reactants including C-reactive protein, D-dimer, and lactate dehydrogenase, as well as absolute neutrophil count. MPO-DNA associated with both cell-free DNA and absolute neutrophil count, while Cit-H3 correlated with platelet levels. Importantly, both cell-free DNA and MPO-DNA were higher in hospitalized patients receiving mechanical ventilation as compared with hospitalized patients breathing room air. Finally, sera from individuals with COVID-19 triggered NET release from control neutrophils *in vitro*. In summary, these data reveal high levels of NETs in many patients with COVID-19, where they may contribute to cytokine release and respiratory failure. Future studies should investigate the predictive power of circulating NETs in longitudinal cohorts, and determine the extent to which NETs may be novel therapeutic targets in severe COVID-19.

## INTRODUCTION

To date, the coronavirus disease 2019 (**COVID-19**) pandemic has affected more than one million individuals from over 250 countries, and has resulted in unprecedented health, social, and economic crises (https://coronavirus.jhu.edu/map.html). The disease is caused by severe acute respiratory syndrome coronavirus 2 (**SARS-CoV-2**), manifesting with flu-like symptoms and a viral pneumonia that progresses to acute respiratory distress syndrome (**ARDS**) and even multi-organ failure in some individuals (1).

Elevated levels of blood neutrophils are an early indicator of SARS-CoV-2 infection, predicting severe respiratory disease and worse outcomes (2, 3). Over the past decade, our group and many others have revealed a pathogenic role for neutrophil-derived neutrophil extracellular traps (**NETs**) in various thrombo-inflammatory states including sepsis (4, 5), thrombosis (6-8), and respiratory failure (9, 10). NETs are extracellular webs of DNA, histones, microbicidal proteins, and oxidant enzymes that are released by neutrophils to corral infections; however, when not properly regulated, NETs have potential to initiate and propagate inflammation and thrombosis (11, 12). Indeed, inhibition of neutrophils and NETs are protective in various models of influenza-associated ARDS (13-16). Although it has yet to be assessed whether NETs contribute to the inflammatory storm that leads to respiratory failure in many COVID-19 patients, there is emerging evidence to implicate inflammatory cytokines such as interleukin (IL)-1β and IL-6 in the COVID-19 milieu (17). Not surprisingly, NETs are intimately intertwined with both cytokines, and especially IL-1β, in many pulmonary and cardiovascular diseases (8, 18-23).

While the definitive accounting of COVID-19 pathophysiology will likely await the development of model systems, it is clear that other pandemic viruses including influenza H1N1, SARS-CoV, and MERS-CoV are associated with neutrophilic infiltration at sites of infection and development of ARDS (24, 25). The acute, exudative phase of ARDS is characterized by an exuberant immune response productive of pro-inflammatory cytokines and chemokines; increased neutrophil infiltration and accumulation in the alveoli; and disruption of the alveolar epithelial-capillary barrier (26). Culturing neutrophils *in vitro* with influenza-infected lung epithelial cells triggers NETosis and augments endothelial damage by culture supernatants (13). Neutrophil-depleted mice demonstrate milder lung pathology in response to influenza infection, including lower levels of thrombomodulin, matrix metalloproteinases, and myeloperoxidase (**MPO**) in bronchoalveolar lavage fluid (13). At the same time, influenza-infected mice are protected by strategies that prevent NETosis including inhibition of superoxide dismutase (13) and MPO (13, 27). In patients with influenza A infection, high levels of NETs predict a poor prognosis (28).

Work to date exploring pathophysiology of COVID-19 has focused especially on macrophages and epithelial cells, with little attention paid to neutrophils and their catalysts, checkpoints, and effector mechanisms—all of which could add actionable context to our understanding of the COVID-19 inflammatory storm. Here, as a first step toward assessing the role of NETs in COVID-19, we sought to measure various markers of NETs in sera of hospitalized patients and to determine their relationship to severity of illness.

## RESULTS

### Detection of NETs in sera of COVID-19 patients

Serum samples were obtained from 50 patients hospitalized with COVID-19 at a large academic hospital (**Table 1**). As compared with serum samples from 30 healthy controls, the COVID-19 samples showed higher levels of cell-free DNA (**Figure 1A**), myeloperoxidase-DNA complexes (**MPO-DNA, Figure 1B**), and citrullinated histone H3 (**Cit-H3, Figure 1C**). The latter two markers are generally regarded as highly specific for NET remnants. While cell-free DNA and MPO-DNA demonstrated a significant correlative relationship (**Figure 1D**), the association between cell-free DNA and Cit-H3 was not significant (**Figure 1E**). For a subset of the patients (n=22), longitudinal serum samples were available. For most, levels of NET markers were relatively stable over several days (**Supplementary Figure 1**). All available samples (n=84) were included in the subsequent correlation analyses. In summary, three markers indicative of NET remnants are elevated in sera of COVID-19 patients as compared with controls.

**Table 1:**
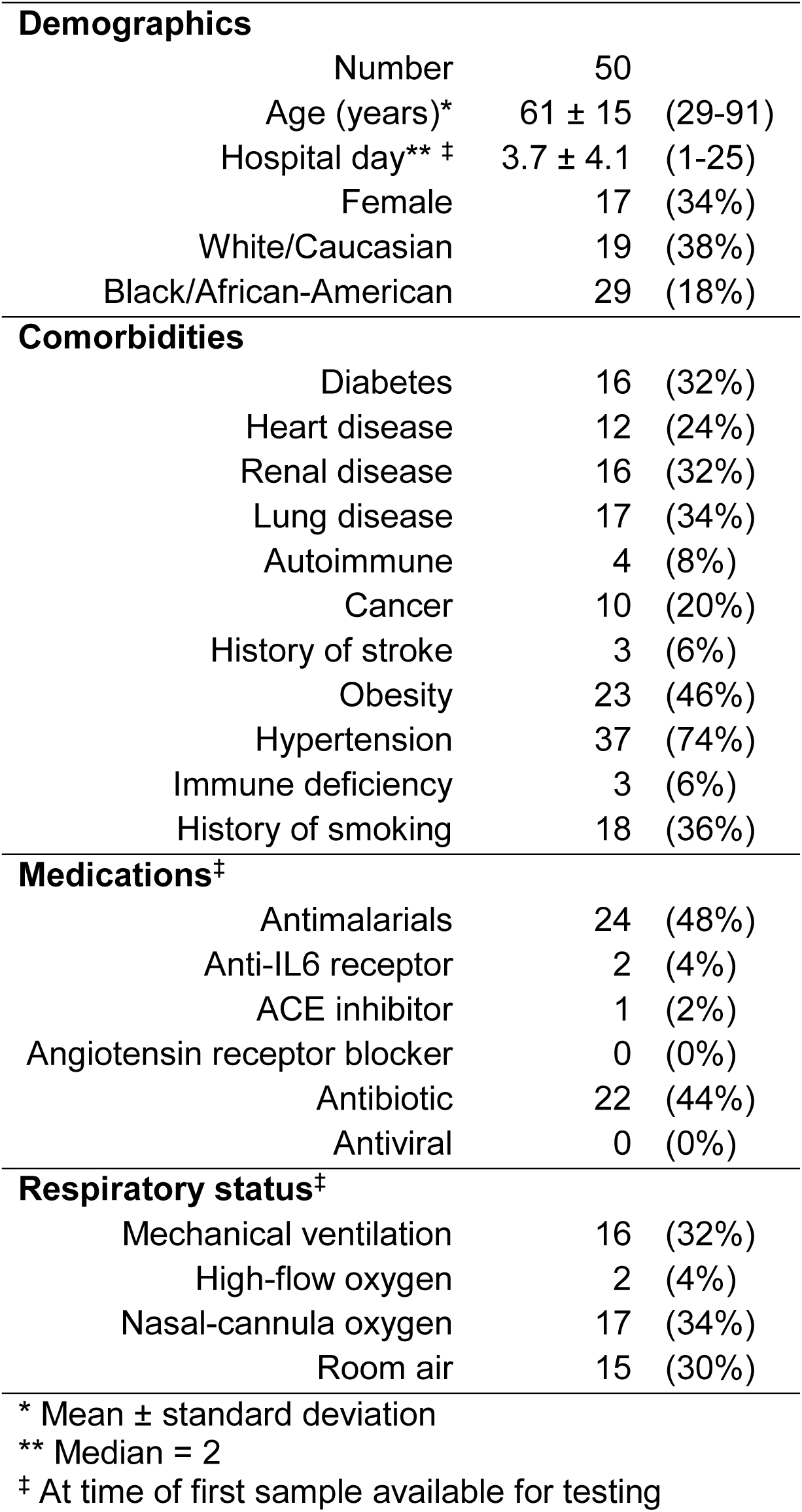
COVID-19 patient characteristics.

**Figure 1:**
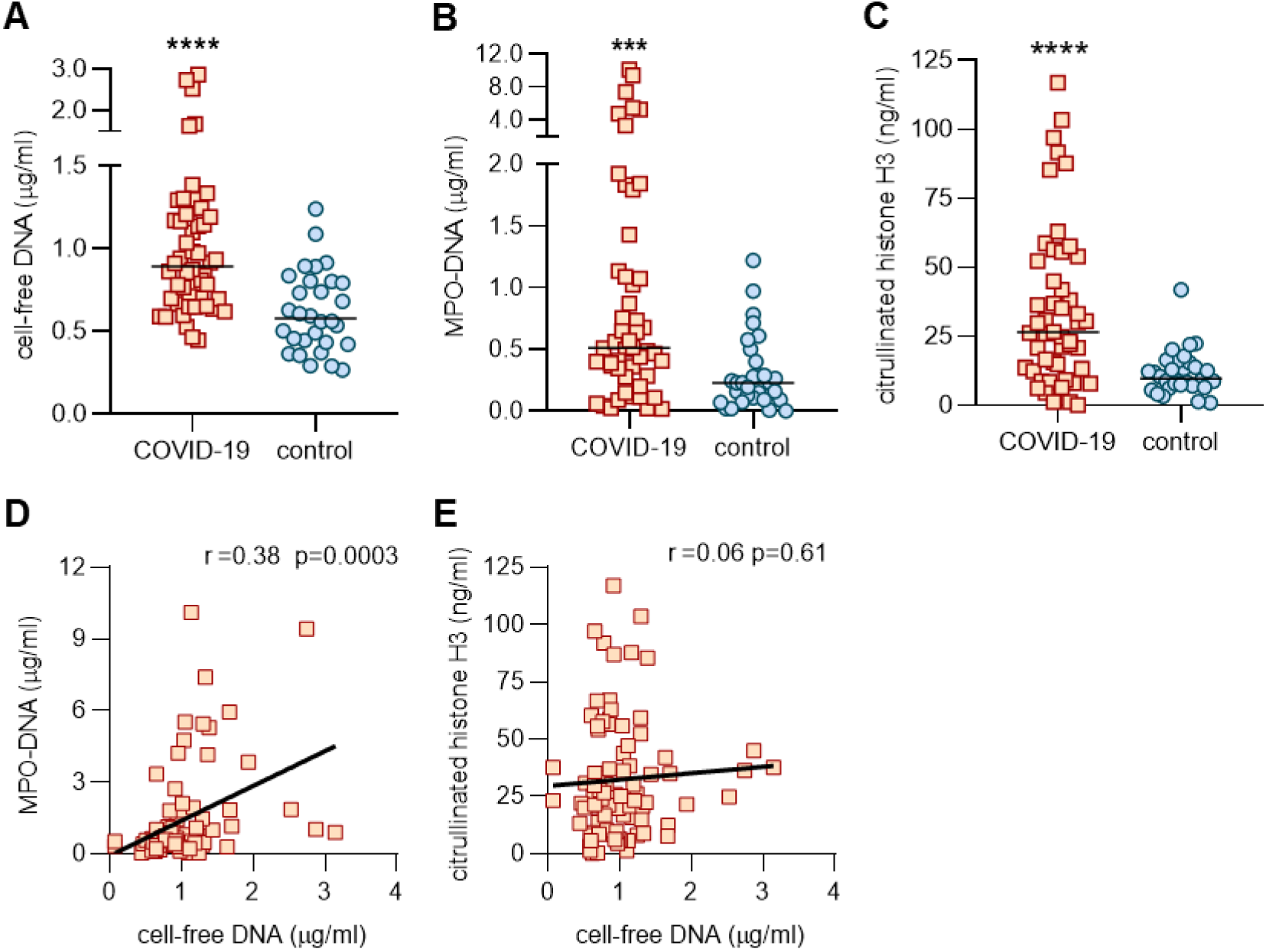
Detection of NETs in sera of COVID-19 patients. Sera from COVID-19 patients (n=50) and healthy controls (n=30) were assessed for cell-free DNA (**A**), myeloperoxidase (MPO)-DNA complexes (**B**), or citrullinated histone H3 (**C**). COVID-19 samples were compared to controls by Mann-Whitney test; ***p<0.001, ****p<0.0001. For the COVID-19 samples, correlation of cell-free DNA with MPO-DNA (**D**) and citrullinated histone H3 (**E**) were assessed. Pearson correlation coefficients were calculated and are shown in the panels.

### Association between NETs and clinical biomarkers

Given that circulating NETs may be drivers of, or form in response to, other blood products, we next asked how the aforementioned NET markers compared to several commonly available clinical tests. Specifically, we assessed potential correlations with C-reactive protein, D-dimer, lactate dehydrogenase, absolute neutrophil count, and platelet count. To draw integral comparisons, we limited the analysis of clinical laboratory measurements to those performed on the same day as serum used for NET assays. Cell-free DNA demonstrated a strong positive correlation with all of the clinical tests other than platelet count (**Figure 2A-D**). When we compared the clinical labs with MPO-DNA, we detected a strong positive correlation with absolute neutrophil count (**Figure 2E**), while C-reactive protein (r=0.10, p=0.44), D-dimer (r=0.05, p=0.72) and lactate dehydrogenase (r=0.17, p=0.22) demonstrated positive slopes that were not statistically significant. Interestingly, Cit-H3 levels were positively correlated with platelet counts (**Figure 2F**), but not the other clinical laboratory measurements. In summary, cell-free DNA and to a lesser extent MPO-DNA and Cit-H3 demonstrate significant correlations with clinical studies routinely used in the care of COVID-19 patients.

**Figure 2:**
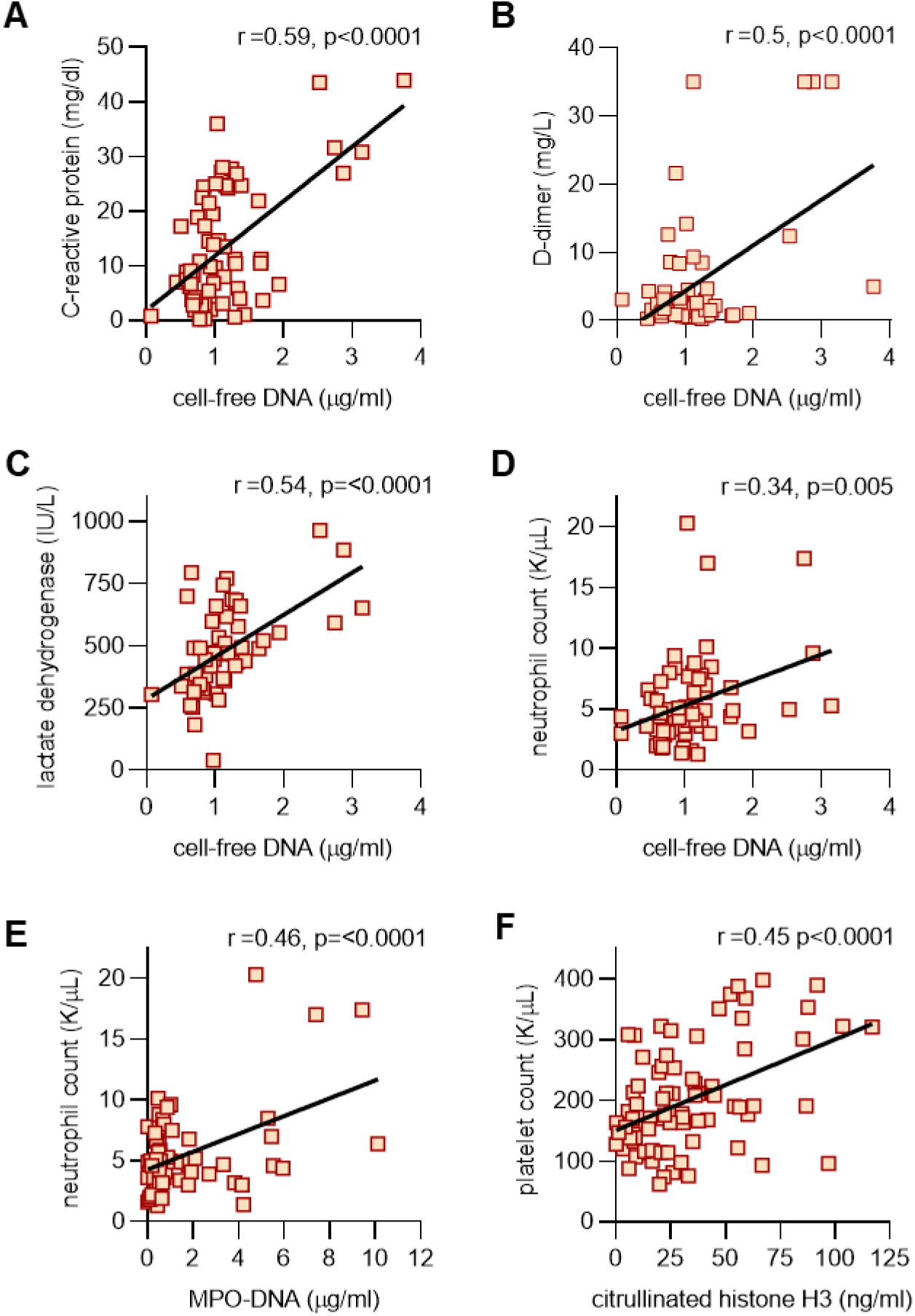
Association between NETs and clinical biomarkers in all available serum samples. Cell-free DNA was compared to clinical laboratory results (when available on the same day), and correlation coefficients were calculated for C-reactive protein (**A**), D-dimer (**B**), lactate dehydrogenase (**C**), and absolute neutrophil count (**D**). In panel **E**, MPO-DNA was compared to absolute neutrophils count and in **F**, citrullinated histone H3 was compared to platelet count. The results of other relevant comparisons are discussed in the text.

### NETs associate with severe disease including mechanical ventilation

We next determined the clinical status associated with each available serum sample. We compared samples from patients with severe COVID-19 (those requiring mechanical ventilation, n=27 samples) to patients with mild or moderate COVID-19 (oxygen saturation >94% on ambient air, n=24 samples). As compared with patients breathing room air, patients requiring mechanical ventilation had significantly higher levels of cell-free DNA (**Figure 3A**) and MPO-DNA (**Figure 3B**), but not Cit-H3 (**Figure 3C**). Absolute neutrophil counts were not significantly higher in the ventilated patients (**Figure 3D**). In summary, these data suggest a possible relationship between level of serum NETs and severity of COVID-19.

**Figure 3:**
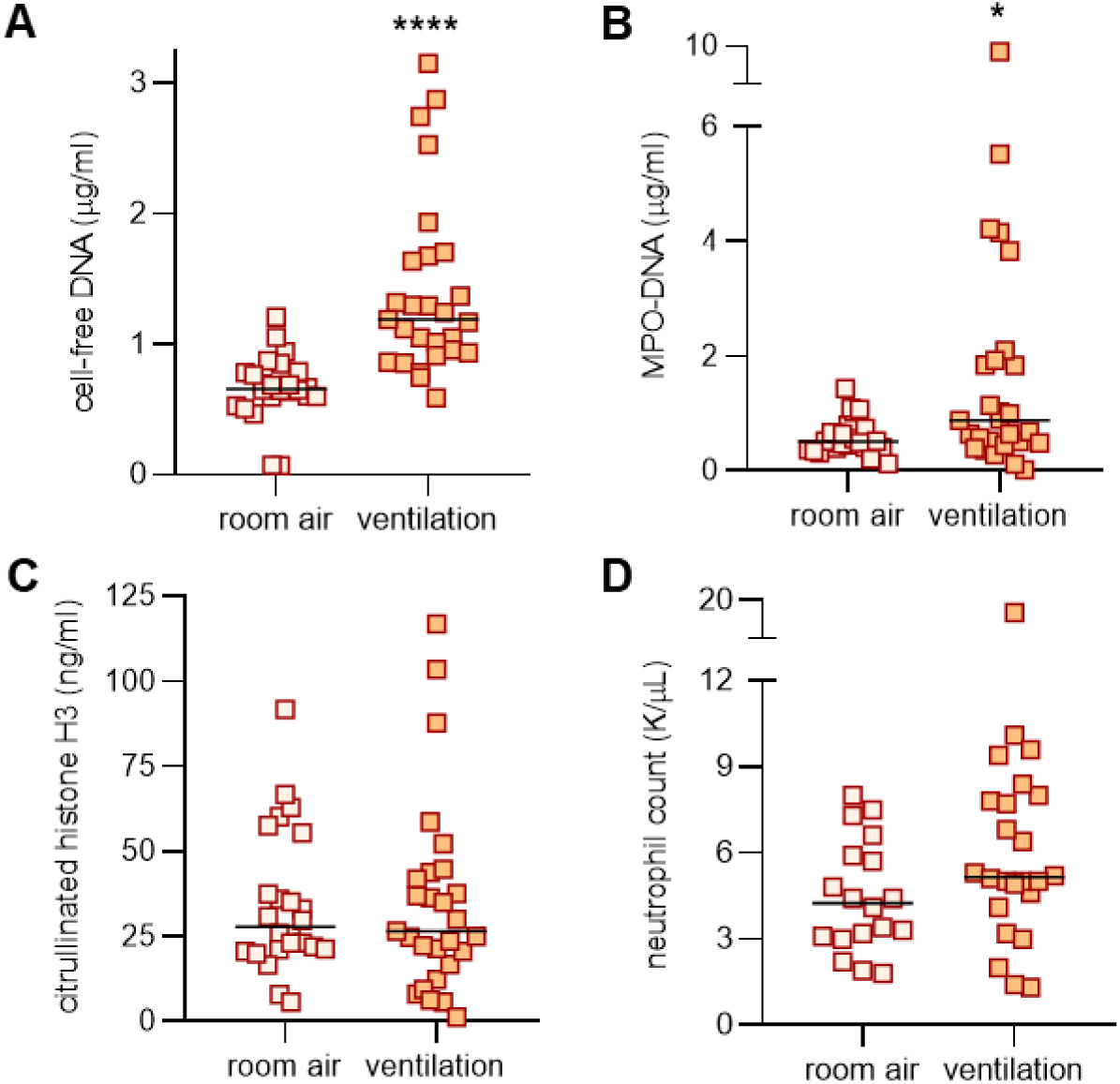
Levels of NETs associate with mechanical ventilation in all available serum samples. Serum samples were grouped by clinical status (room air versus mechanical ventilation), and analyzed for cell-free DNA (**A**), MPO-DNA (**B**), citrullinated histone H3 (**C**), and absolute neutrophil count (**D**). Groups were compared by Mann-Whitney test; ****p<0.0001, *p<0.05. For panel D, the p value was 0.08.

### COVID-19 sera trigger control neutrophils to release NETs

If COVID-19 presents a milieu favoring NETosis, we reasoned that direct exposure of control neutrophils to patient sera (without any additional agonist) would trigger these healthy neutrophils to undergo NETosis. As compared with heterologous control sera, the patient sera robustly promoted NETosis whether measured by externalization of DNA (**Figure 4A**) or release of DNA-bound MPO enzyme (**Figure 4B**). Immunofluorescence microscopy demonstrated extracellular chromatin structures decorated with neutrophil elastase characteristic of NETs (**Figure 4C**). In summary, serum samples from COVID-19 patients robustly trigger healthy neutrophils to undergo NETosis.

**Figure 4:**
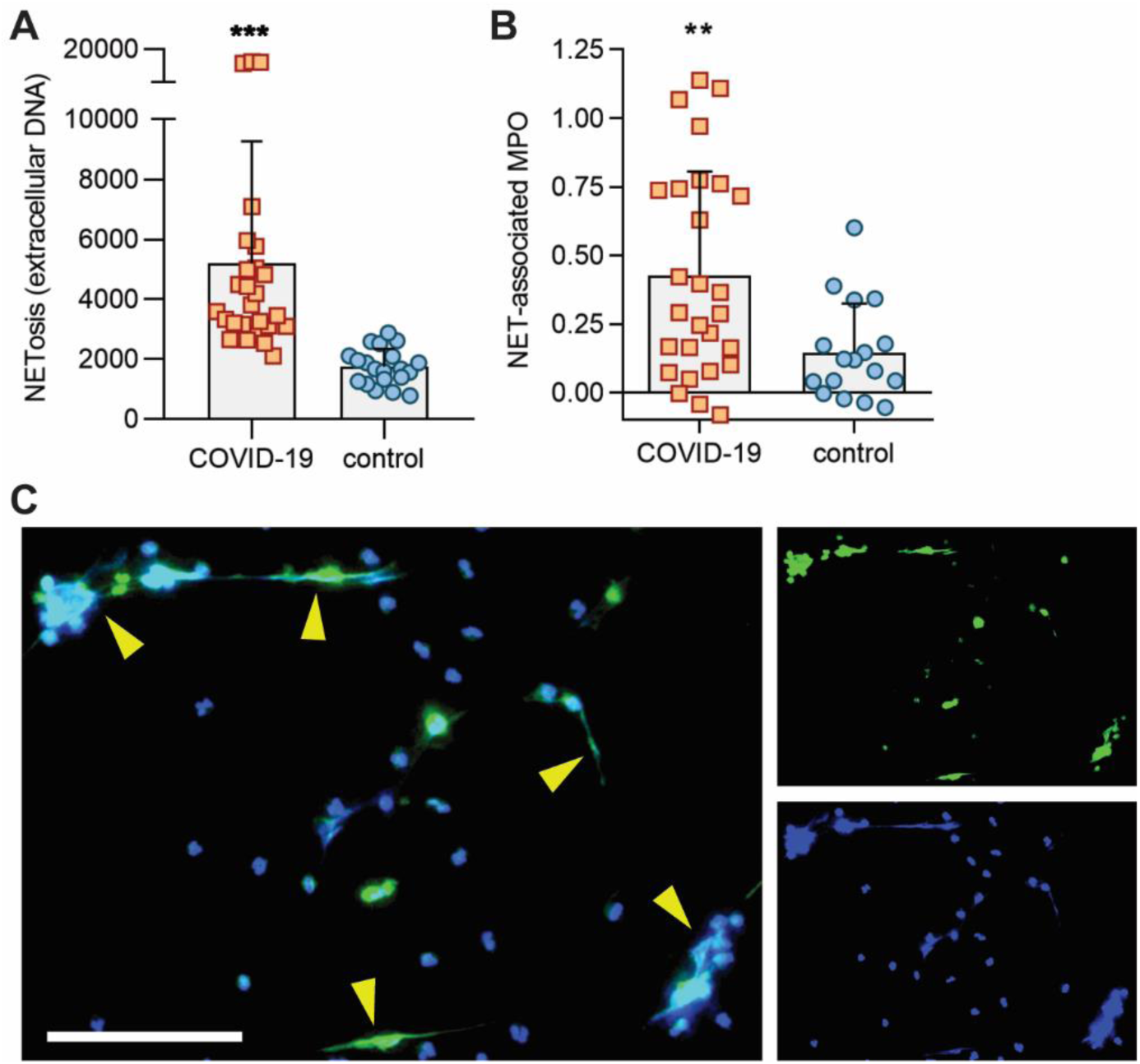
COVID-19 sera trigger control neutrophils to release NETs. COVID-19 samples (for which sufficient sera were available) were tested for their ability to trigger neutrophils isolated from healthy controls to undergo NETosis. **A**, NETosis was quantified using the cell-impermeant dye SYTOX Green as described in Methods. Fluorescence intensity (excitation/emission 504/523) is shown on the y-axis. Bars demonstrate mean and standard deviation while each data point represents a unique patient/control; ***p<0.001 by t test. **B**, In an independent set of experiments, NETosis was quantified as nuclease-liberated myeloperoxidase (MPO) activity. Absorbance at 450 nm is shown on the y-axis after subtracting background from untreated cells. Bars demonstrate mean and standard deviation while each data point represents a unique patient/control; **p<0.01 by t test. **C**, Representative image of control neutrophils cultured with 10% COVID-19 serum. Neutrophil elastase is stained green and DNA is stained blue. Scale bar=100 microns. The yellow arrows highlight some examples of NET strands.

## DISCUSSION

Here, we report for the first time elevated levels of serum NETs in many hospitalized patients with COVID-19. We measured three markers commonly used to detect NET remnants in blood (cell-free DNA, Cit-H3, and MPO-DNA), and found significant elevations in all three. We also found that COVID-19 sera are potent stimulators of NETosis when added to control neutrophils. Together, these data provide evidence that COVID-19, at least in hospitalized patients, is a pro-NETotic state. The triggers of NETosis in COVID-19 are potentially myriad and will require further investigation. Possibilities include virus-damaged epithelial cells (13, 29), activated platelets (30, 31), activated endothelial cells (32), and inflammatory cytokines such as IL-1β (19, 33), IL-8 (34, 35), granulocyte colony stimulating factor (36, 37), and likely others (17).

Of the markers we tested, cell-free DNA was most closely associated with traditional inflammatory markers used to track COVID-19 including C-reactive protein, D-dimer, and lactate dehydrogenase. Notably, although cell-free DNA is not a highly specific marker for NETs, it was strongly correlated with absolute neutrophil count, as was the more specific marker of NETs, MPO-DNA. Somewhat unexpectedly, Cit-H3 did not correlate well with the other two markers, but did associate strongly with platelet levels. It is believed that the predominant driver of histone citrullination (i.e., production of Cit-H3) in NETs is the enzyme peptidlyarginine deiminase 4 (**PAD4**) (38). However, neutrophils can be triggered to undergo NETosis by a variety of stimuli, and *in vitro* studies demonstrate that not all pathways to NETosis are equally reliant on PAD4 activity (39); for example, stimuli that lead to robust reactive oxygen species production may be relatively PAD4-independent (40). The dichotomy between MPO-DNA and Cit-H3 levels in the COVID-19 sera tested here potentially suggests that two or more pathways to NETosis are active in COVID-19 patients, with the pathway leading to Cit-H3 perhaps having some relationship to platelets (41).

NETs were first described in 2004 as a novel pathogen eradication strategy that could function as an alternative to phagocytosis (35), but it is now recognized that NETs have double-edged-sword properties and likely exacerbate (and in some cases even initiate) autoimmune and vascular diseases (42). NETs present and stabilize a variety of oxidant enzymes in the extracellular space, including MPO, NADPH oxidase, and nitric oxide synthase (43), while also serving as a source of extracellular histones that carry significant cytotoxic potential (44, 45). In light of these toxic cargo, it is not surprising that NETs play a role in a variety of lung diseases including cystic fibrosis (where they occlude larger airways) (46), smoking-related lung disease (47), and, with particular relevance here, pathogen-induced acute lung injury and ARDS (13, 48, 49). NETs have also been very well studied in the setting of cardiovascular disease where they infiltrate and propagate inflammation in the vessel wall (50), and, when formed intravascularly, occlude arteries (51), veins (52), and microscopic vessels (53). Early studies of COVID-19 suggest a high risk of morbid arterial events (54), and one can speculate that the risk of venous thrombosis will increasingly reveal itself as more data become available (55).

Severe COVID-19 appears to be defined by neutrophilia, as well as elevations in IL-1β, IL-6, and D-dimer (17), the latter suggesting hyperactivity of the coagulation system. All these findings have significant potential for cross-talk with NETs. NETs are linked to IL-1β (both upstream and downstream) in cardiovascular and pulmonary diseases (18-21), including as described by our group for venous thrombosis (8). The same is true for IL-6, either directly (22), or perhaps with IL-1β as an intermediary (23). Of course, as discussed above, examples of NETs as drivers of thrombosis are myriad, as intravascular NETosis is responsible for initiation and accretion of thrombotic events in arteries, veins, and—particularly pertinent to COVID-19— the microvasculature, where thrombotic disease can drive end-organ damage in lungs, heart, kidneys, and other organs (56, 57). Mechanistically, NETs, via electrostatic interactions, activate the contact pathway of coagulation (58), while also presenting tissue factor to activate the intrinsic pathway (59). Simultaneously, serine proteases in NETs dismantle natural brakes on coagulation such as tissue factor pathway inhibitor and antithrombin (60). Bidirectional interplay between NETs and platelets may also be critical for COVID-19-associated microvascular thrombosis as has been characterized in a variety of disease models (57, 58).

Of interest, a recent small study performed in China suggested potential efficacy of the adenosine-receptor agonist, dipyridamole in severe cases of COVID-19 (61). Dipyridamole is an FDA-approved drug that our group recently discovered to inhibit NET formation by activation of adenosine A2A receptors (6). In the aforementioned trial, patients with COVID-19-associated bilateral pneumonia were treated with oral dipyridamole for seven days, in additional to treatment with antiviral agents (61). As compared with controls, dipyridamole-treated patients demonstrated improvements in platelet counts and D-dimer levels (61). Given the urgent need for effective treatments of COVID-19, a randomized study to characterize the impact of dipyridamole on COVID-19-related NETosis, thrombo-inflammatory storm, and, of course, outcomes may be warranted.

Other approaches to combatting NETs have been reviewed (62, 63), and include the dismantling of already-formed NETs (deoxyribonucleases) and strategies that might prevent initiation of NET release, including inhibitors of neutrophil elastase and PAD4. While our study certainly has limitations including the use of serum (plasma is typically preferred by our group and others for measurement of NETs) and a relatively small sample size, we hope it will ignite further research into the role of neutrophil effector functions in the complications of COVID-19. As a first step, future studies should investigate the predictive power of circulating NETs in well-phenotyped longitudinal cohorts. Furthermore, given the dichotomy we found here between MPO-DNA and Cit-H3, investigators should be encouraged to continue to include diverse markers of NETosis in future studies. As we await definitive antiviral and immunologic solutions to the current pandemic, we posit that anti-neutrophil therapies may be part of a personalized strategy for some individuals affected by COVID-19 who are at risk for progression to respiratory failure.

## METHODS

### Human samples

Serum samples from 50 hospitalized COVID-19 patients (84 total samples) were used in this study. Blood was collected into serum separator tubes containing clot activator and serum separator gel by a trained hospital phlebotomist. After completion of biochemical testing ordered by the clinician, the remaining serum was stored at 4°C for up to 48 hours before it was deemed “discarded” and released to the research laboratory. Serum samples were immediately divided into small aliquots and stored at -80°C until the time of testing. All 50 patients had a confirmed COVID-19 diagnosis based on FDA-approved RNA testing. This study complied with all relevant ethical regulations, and was approved by the University of Michigan Institutional Review Board (HUM00179409), which waived the requirement for informed consent given the discarded nature of the samples. Healthy volunteers were recruited through a posted flyer; exclusion criteria for these controls included history of a systemic autoimmune disease, active infection, and pregnancy. For preparation of control serum, blood was collected into serum separator tubes containing clot activator and serum separator gel by a trained hospital phlebotomist. These serum samples were divided into small aliquots and stored at -80°C until the time of testing.

### Quantification of cell-free DNA

Cell-free DNA was quantified in sera using the Quant-iT PicoGreen dsDNA Assay Kit (Invitrogen, P11496) according to the manufacturer’s instructions.

### Quantification of citrullinated histone H3

Citrullinated histone H3 was quantified in sera using the Citrullinated Histone H3 (Clone 11D3) ELISA Kit (Cayman, 501620) according to the manufacturer’s instructions.

### Quantification of MPO-DNA complexes

MPO-DNA complexes were quantified similarly to what has been previously described (64). This protocol used several reagents from the Cell Death Detection ELISA kit (Roche). First, a high-binding EIA/RIA 96-well plate (Costar) was coated overnight at 4°C with anti-human MPO antibody (Bio-Rad 0400-0002), diluted to a concentration of 1 µg/ml in coating buffer (Cell Death kit). The plate was washed two times with wash buffer (0.05% Tween 20 in PBS), and then blocked with 4% bovine serum albumin in PBS (supplemented with 0.05% Tween 20) for 2 hours at room temperature. The plate was again washed five times, before incubating for 90 minutes at room temperature with 10% serum or plasma in the aforementioned blocking buffer (without Tween 20). The plate was washed five times, and then incubated for 90 minutes at room temperature with 10x anti-DNA antibody (HRP-conjugated; from the Cell Death kit) diluted 1:100 in blocking buffer. After five more washes, the plate was developed with 3,3’,5,5’-Tetramethylbenzidine (TMB) substrate (Invitrogen) followed by a 2N sulfuric acid stop solution. Absorbance was measured at a wavelength of 450 nm using a Cytation 5 Cell Imaging Multi-Mode Reader (BioTek). Data were normalized to *in vitro*-prepared NET standards included on every plate, which were quantified based on their DNA content.

### Human neutrophil purification

For neutrophil preparation, blood from healthy volunteers was collected into sodium citrate tubes by standard phlebotomy techniques. The anticoagulated blood was then fractionated by density-gradient centrifugation using Ficoll-Paque Plus (GE Healthcare). Neutrophils were further purified by dextran sedimentation of the red blood cell layer, before lysing residual red blood cells with 0.2% sodium chloride. Neutrophil preparations were at least 95% pure as confirmed by nuclear morphology.

### NETosis assay (SYTOX Green)

A cell-impermeant dye SYTOX Green (Thermo Fisher) was used to measure NETosis. Briefly, purified neutrophils were resuspended in 1x PBS (Gibco). 1×10^5^ neutrophils were seeded into each well of a 0.001% poly-L-lysine (Sigma)-coated 96-well black clear-bottom non-tissue culture plate (Costar), and were allowed to adhere for 20 minutes at 37°C and 5% CO2. PBS was gently removed and control/patient serum (diluted to 10% in RPMI culture media supplemented with L-glutamine) was carefully added without disrupting adherent cells. SYTOX Green was added at the same time to a final concentration of 500 nM. All treatments were done in triplicate. Cells were allowed to undergo NETosis for 4 hours. Culture media was then gently removed and fresh 1x PBS was added to each well. Fluorescence was quantified at excitation and emission wavelengths of 504 nm and 523 nm, respectively, using a Cytation 5 Cell Imaging Multi-Mode Reader (BioTek). Data were collected using the area-scan setting of the plate reader.

### NETosis assay (NET-associated MPO)

Purified neutrophils were resuspended in RPMI media (Gibco) supplemented with 0.5% bovine serum albumin (BSA, Sigma), 0.5% heat-inactivated fetal bovine serum (FBS, Gibco) and L-glutamine. Neutrophils (1 × 10^5^ /well) were cultured in a 96-well tissue culture plate (Costar) in the presence of either patient or control serum, diluted to a final concentration of 10%. Plates were incubated for 3 hours at 37°C and 5% CO2. To collect NET-associated MPO, the culture media was discarded (to remove any soluble MPO) and replaced with 100 µL of PBS supplemented with 5 U/ml Micrococcal nuclease (Thermo Fisher). After 10 minutes at 37°C, digestion of NETs was stopped with 10 mM EDTA. Supernatants were transferred to a v-shaped 96 well plate, and centrifuged at 400xg for 5 minutes to remove debris. Supernatants were then transferred into a new flat-bottom 96-well plate. To quantify MPO activity, an equal volume of 3,3’,5,5’-tetramethylbenzidine (TMB) substrate (Thermo Fisher) was added to each well. After 10 minutes of incubation in the dark, the reaction was stopped by the 2N sulfuric acid solution. Absorbance was measured at 450 nm using a Cytation 5 Cell Imaging Multi-Mode Reader.

### NETosis assay (microscopy)

For immunofluorescence microscopy, 1×10^5^ neutrophils were seeded onto coverslips coated with 0.001% poly-L-lysine (Sigma) and cultured as for the above assays. Samples were then fixed with 4% paraformaldehyde for 10 minutes at room temperature, followed by blocking with 10% FBS in PBS. The primary antibody was against neutrophil elastase (Abcam 21595, diluted 1:100), and the FITC-conjugated secondary antibody was from SouthernBiotech (4052-02, diluted 1:250). DNA was stained with Hoechst 33342 (Invitrogen). Images were collected with a Cytation 5 Cell Imaging Multi-Mode Reader.

### Statistical analysis

Normally-distributed data were analyzed by two-sided t test and skewed data were analyzed by Mann-Whitney test. Data analysis was with GraphPad Prism software version 8. Correlations were tested by Pearson’s correlation coefficient. Statistical significance was defined as p<0.05 unless stated otherwise.

## Data Availability

Requested data will be made available in timely fashion once the manuscript is published in a peer-reviewed journal.

## ACKNOWLEDGEMENTS

The work was supported by a COVID-19 Cardiovascular Impact Research Ignitor Grant from the Michigan Medicine Frankel Cardiovascular Center as well as by the A. Alfred Taubman Medical Research Institute. YZ was supported by career development grants from the Rheumatology Research Foundation and APS ACTION. JAM was partially supported by the VA Healthcare System. YK was supported by the NIH (K08HL131993, R01HL150392), Falk Medical Research Trust Catalyst Award, and the JOBST-American Venous Forum Award. JSK was supported by grants from the NIH (R01HL115138), Lupus Research Alliance, and Burroughs Wellcome Fund. YZ and JSK also thank all members of the “COVID-19 NETwork” for their helpful advice and encouragement.

## AUTHORSHIP

YZ, SY, HS, KG, JM, MZ, and CB conducted experiments and analyzed data. YZ, AW, BJB, ME, RJW, YK, and JSK conceived the study and analyzed data. All authors participated in writing the manuscript and gave approval before submission.

**Supplementary Figure 1:**
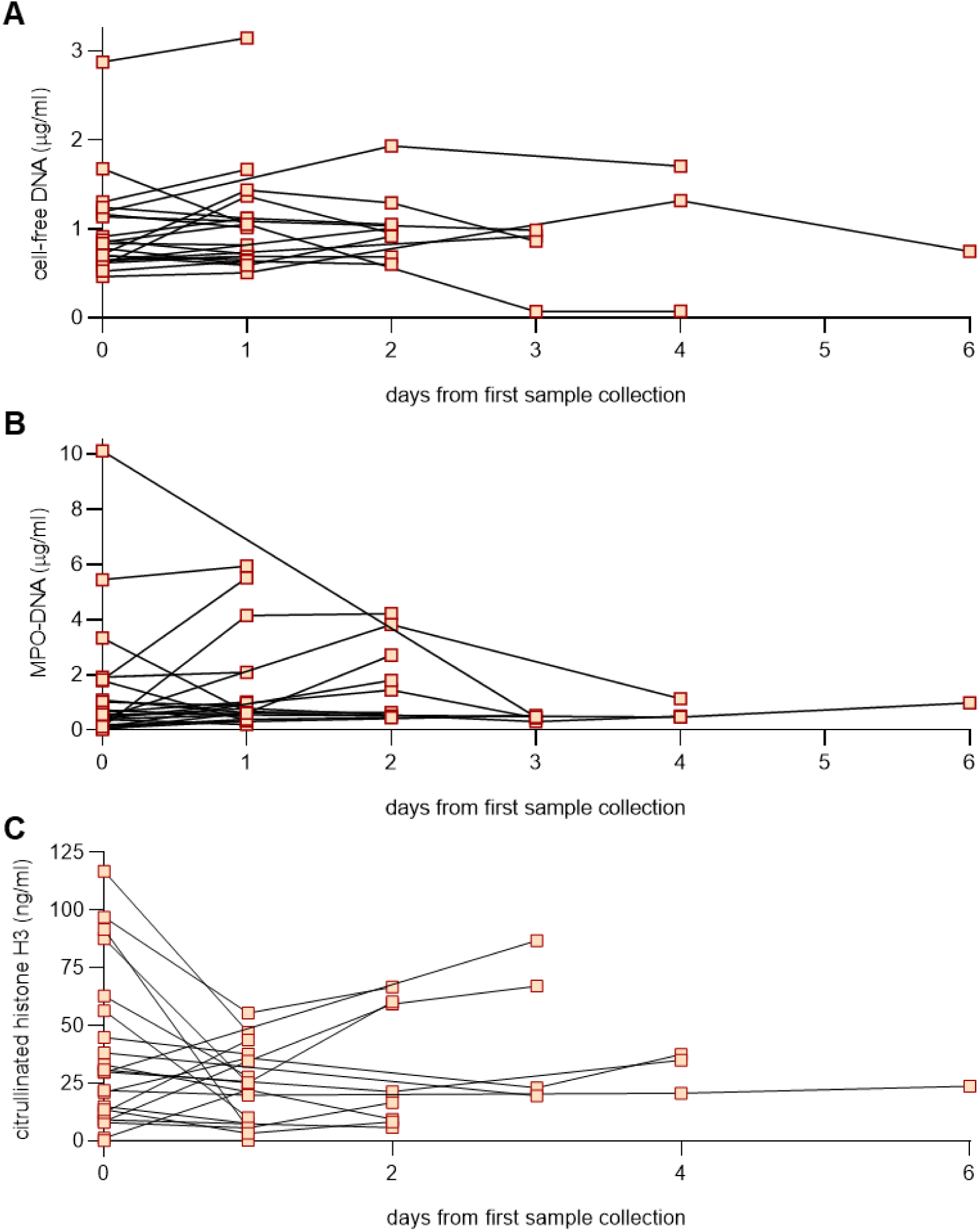
Markers of NETs in longitudinal serum samples. For 22 patients, serum samples were available from multiple days. Data are plotted here for cell-free DNA (**A**), myeloperoxidase (MPO)-DNA (**B**), and citrullinated histone H3 (**C**).

